# Predicting Stress in Teens from Wearable Device Data Using Machine Learning Methods

**DOI:** 10.1101/2020.11.26.20223784

**Authors:** Claire W. Jin, Ame Osotsi, Zita Oravecz

## Abstract

Stress management is a pervasive issue in the modern high schooler’s life. Despite many efforts to support adolescents’ mental well-being, teenagers often fail to recognize signs of high stress and anxiety until their emotions have escalated. Being able to identify early signs of these intense emotional states and predict their onset using physiological signals collected passively in real-time could help teenagers improve their awareness of their emotional well-being and take a more proactive approach to managing their emotions. To evaluate the potential of this approach, we collected data from high schoolers with Empatica E4 wearable health monitors (wristband) while they were living their daily lives. The data consisted of stressful event reports and physiological markers over the course of 4 weeks. We developed a random forest model and a support vector machine model and systematically assessed their performance in terms of predicting the onset of stress events and identifying physiological signals of stress. The models showed strong performance in terms of these measures and provided insights on physiological indicators of adolescent stress.

## 1. Introduction

In recent years, it has become increasingly clear that mental health is a serious concern for modern teenagers. Stress from school, extracurriculars, college, and their social lives have major negative repercussions for adolescents’ physical and mental well being. A 2017 survey [2] conducted in the State College Area School District by the Pennsylvania Department of Education showed that a high number of teenage students in the district reported being at elevated risk for anxiety, depression, low self-efficacy, and suicidal ideation. This problem is not unique to teens in that area, but an epidemic [3] in today’s world. Despite many efforts to support teenagers’ mental well-being, teens often fail to recognize or pay attention to signs of stress until their emotions have escalated, or are unwilling to discuss their feelings with or seek support from others. Being able to identify early signs of high stress, anxiety, and “low” feelings and predict their onset using physiological signals collected passively in real-time could help teens improve their awareness of their emotional well-being and take a more proactive approach to managing their emotions.

As technology has improved rapidly and become increasingly mobile and personalized, access to wearable health monitors, such as FitBits, has greatly improved. Wearable health monitors are relatively unobtrusive tools for collecting physiological data.When combined with predictive models, they can help people proactively handle serious health issues, both physical and mental. Devices such as Empatica E4, sociometric badges, and EEG bands provide high-quality, real-time, ecologically valid physiological and social data streams in real-life contexts, often with minimal burden to participants. Data from wearables have shown promise to help identify psychological states such as stress [9, 10] and emotional arousal [12, 14]. Getting reliable predictions of psychological states from wearable device data would allow for contributing to the design of cost-effective prevention and intervention programs; for example, helping prevent costly negative health outcomes and support the adoption or maintenance of positive health behaviors. Before wearable data can be used to inform intervention, however, well-designed analytical approaches are needed to distill it into actionable decision rules. This study provides new insights into optimally selecting such approaches.

## 2. Data Collection and Preparation

### 2.1 Study Setting

The data in our study was collected from eight participants, all female highschoolers attending a public high school in Pennsylvania. For one participant, data was collected from the beginning of October 2019 to the end of January 2020. For the other seven, data was collected from the beginning of March 2020 to mid April 2020. The participants wore the Empatica E4 wristband [13] on their non-dominant wrist during the study period to collect physiological data and performed their regular life activities. The participants were instructed to press a specific button on the device whenever they felt stressed at the onset of their physical symptoms (e.g. feeling hot, sweating, sound of blood rushing past ears, belly pain, etc.).

Each participant received one device and a charging dock, along with instructions on how to care for the device and upload data. The data was uploaded through the locally downloaded data management portal, E4 Manager application. After logging in with the assigned account, the device was to be connected to the charging piece, which needed to be connected to the computer using a microUSB before data could be uploaded and new firmware for the device could be downloaded using the E4 Manager interface. This data was then visible on the Empatica cloud, E4 Connect, that is accessible through logging in on a website using the same credentials as the account used to upload data through E4 Manager.

Due to its limited battery and storage capacity, the participants were instructed to wear the device continuously for two days, removing the device to recharge and upload data every other night as they sleep. The participants were also instructed to avoid turning the device off during a session if they needed to remove it. Instead, they were to take it off and leave it somewhere safe and dry until they could put it on again.

At the end of the study period, we examined the number of stress events from each individual and dropped two participants with a very low number of stress events (3 and 5), yielding a final sample size of N = 6. The number of stress events ranged from 10 to 33 in the remaining participants.

### 2.2 Passively Collected Physiological Data

Empatica’s E4 contains four primary sensors and an event mark button. A photoplethysmographic (PPG) sensor measures blood volume pulse (BVP) at a rate of 64 Hz (measurements per second), from which cardiovascular information such as heart rate, heart rate variability, and others may be derived. Tonic electrodermal activity (EDA) [15], also called Skin Conductance Response (SCR), is recorded at 4 Hz using a pair of stainless steel electrodes mounted on the underside of the band, which reports electrical properties of the skin that have been shown to be impacted by stress. A 3-axis accelerometer captures motion of the wrist at a sampling rate of 32 Hz. We derive an ACC “data stream” (time series sequence from one sensor) using root mean square acceleration over the x, y and z coordinates in the accelerometer data stream. The E4 is also equipped with infrared thermopile reading peripheral skin temperature at 4 Hz. Finally, it has an event mark button for the participant to press that marks the data at that time with an “event.” In this study, such an event indicated the onset of noticeable physical stress symptoms. The unique combination of available information from these sensors and the event mark button allow us to leverage the information of multiple data streams: several different physical properties, other properties that can be derived from them, and input from the wearer. Figure 1 below shows how the raw data from the sensors is displayed in the E4 Connect Portal.

**Figure 1:**
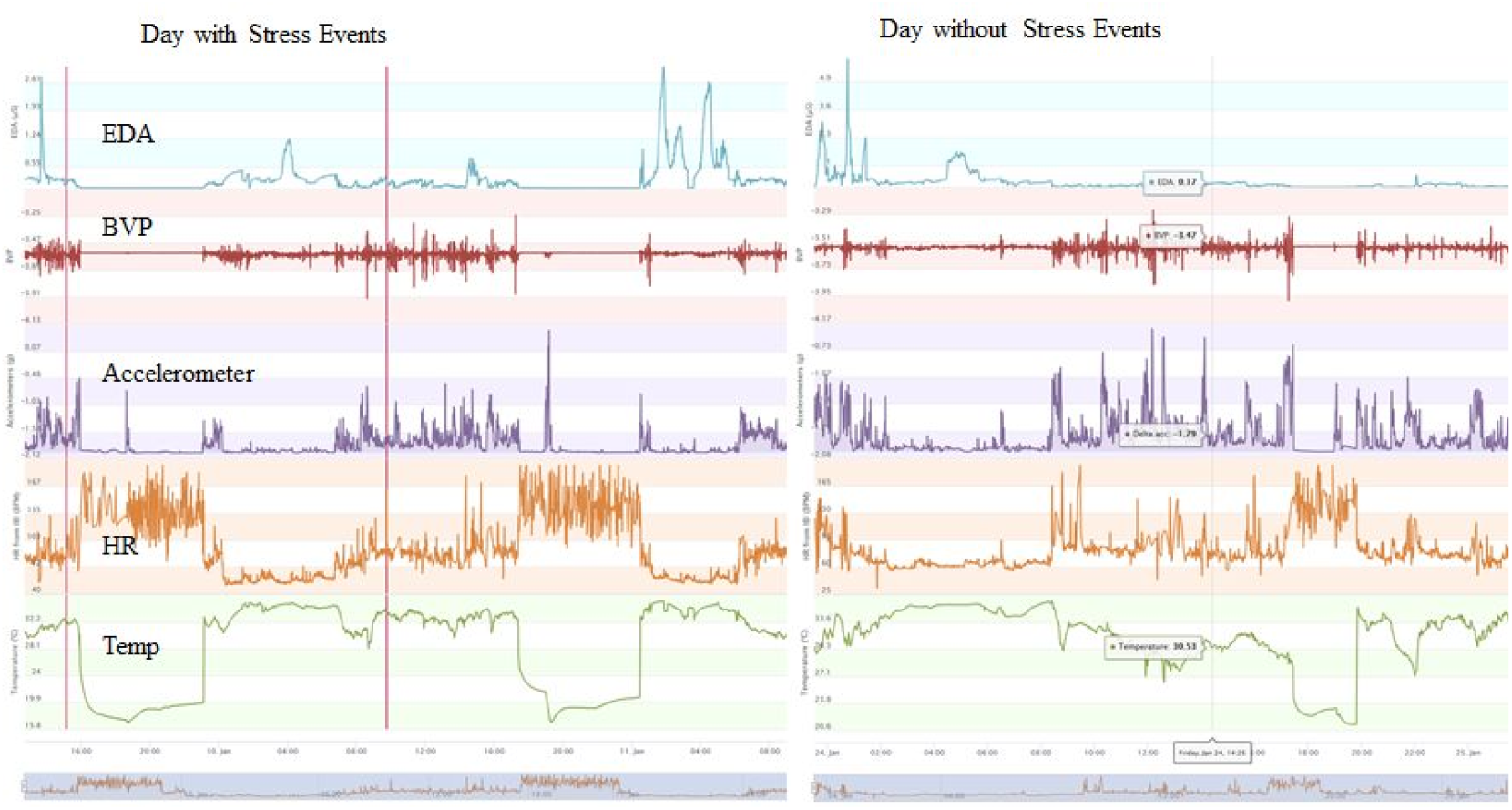
Time series of raw BVP, EDA, accelerometer, temperature, and HR data for a day with stress events (left) and a day without stress events (right). The red vertical lines in the left figure mark button presses.

**Figure 2:**
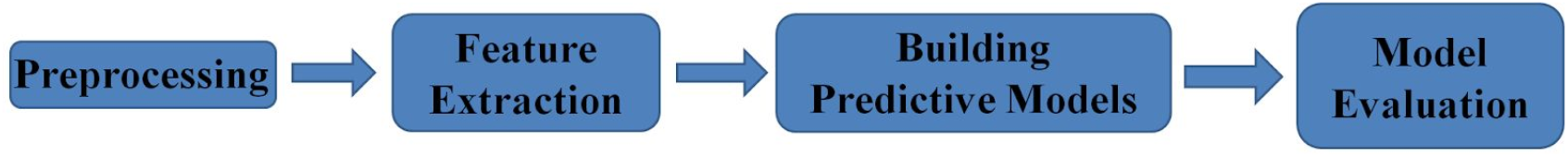
Our data analysis workflow

### 2.3 Data Preprocessing with R and python

Before the data from the sensors could be used in a classifier, it needed to be preprocessed. The main part of this process was downsampling. The sensors on the E4 wristband report measurements at different speeds. The PPG sensor reports BVP at 64 Hz, the accelerometer reports at 32 Hz, while EDA and temperature are measured at 4 Hz. We downsampled the raw data streams to 1 Hz (one measurement every second) by computing the mean of the raw data in that interval. This was done to both reduce computing time, as we aim at developing models that can do timely computations for near real-time inference, and homogenize the frequency of the data for each metric. Although there is no clear theoretical reasoning for the specific choice of 1 second windows, we found that more frequent sampling greatly increases memory requirements and computational time without much corresponding increase in accuracy. Pilot testing showed the sampling frequency of 1 Hz has minimal impact on the accuracy of prediction of a reported stress event.

The measurements for each metric used in our model (BVP, EDA, Temp, ACC, and HR) from the 15 minutes prior to each button press were then extracted to form an *n* by 900 matrix, where *n* is the total number of events during the collection period and 900 is the number of measurement collected in 15 minutes (1 measurement per second, 15 × 60 = 900) for each metric. Then the same *n* number of non-events were chosen randomly and were processed in the same way. In doing so, each individual has balanced data and we maximize the number of events used for training the models. The event matrix and non-event matrix for each metric were then combined to form a 2*n* × 900 data matrix.

To ensure that the non-events are not correlated with the events, the non-events were randomly chosen from times far away from the events using the locator() function in R. Several different sets of randomly chosen non-events were used to fit the classifier for the participant who collected data from October through January to test its flexibility, as shown in the results section (Table 1).

**Table 1:** The AUCs for the overall classifier and the single-metric classifiers for four different data sets from a single person (non-events chosen were varied) is shown here along with the average AUC and standard deviation for each type of classifier.

| Different sets of non-events | Classifier AUC |  |  |  |  |
| --- | --- | --- | --- | --- | --- |
|  | Overall | BVP only | EDA only | Temp only | HR only |
| a | 0.894 | 0.842 | 0.686 | 0.697 | 0.875 |
| b | 0.892 | 0.747 | 0.797 | 0.839 | 0.666 |
| c | 0.827 | 0.774 | 0.677 | 0.571 | 0.780 |
| d | 0.833 | 0.874 | 0.747 | 0.793 | 0.786 |
| Average | 0.862 | 0.809 | 0.727 | 0.725 | 0.777 |
| Standard deviation | 0.036 | 0.059 | 0.056 | 0.118 | 0.086 |

## 3. Machine Learning Approach to Predicting Stress Events

### 3.1 Feature-based Binary Classification

#### 3.1.1 Feature extraction

In order to study which characteristics of a time series are important for stress classification, we first extracted features of the time-series. These features may represent either global (e.g. grand mean, the maximum/minimum of the time series) or local characteristics of the time series structured around a specific time point (e.g. a wavelet transform coefficient). We used the python package *tsfresh* to extract features for each metric. These features range from simple measures such as minimum, maximum, variance, and autocorrelation to count-based measures such as the number of measurements above or below the mean to more intricate features such as approximate entropy, frequency domain measures such as power spectrum density, and time-frequency measures such as wavelet transform coefficients. See the *tsfresh* documentation for details [1].

As explained, for each dataset, a 2*n* × 900 matrix was generated for each metric. These matrices were then fed into *tsfresh* separately to return the values of 756 metric-associated features [7] for the 15 minute interval before each selected time point (event or non-event), resulting in a matrix of n by 756 features as columns. The matrices associated with each set of non-events were then combined, resulting in 756 × 6 = 4536 total features for each data set fed into *tsfresh* since 6 metrics (BVP, EDA, Temp, ACC - root mean squared and root mean squared velocity, and HR) were considered in our model. The extracted features were used as the input for the classifier.

#### 3.1.2 Classification models

We used random forest (RF) to classify the features extracted from the physiological signals [5]. Random forest is an ensemble classifier, consisting of a large number of decision trees. Each decision tree is a simplistic classifier which attempts to find a series of simple binary conditions that are most predictive of a given outcome. The predictions of the many decision trees in the random forest are combined using majority vote. By combining a series of these conditions into a tree, it is possible to capture arbitrarily complex interactions among the predictors. However, because the selection of decision criteria is performed by searching a large number of features and potential split points, it has a tendency to overfit data sets. To reduce the tendency of decision trees to overfit, randomness is introduced into both the data selection and feature selection. As a result, random forests do not require much fine-tuning of parameters and are also robust to the presence of noise in the predictors.

Random forests also provide an inherent measure of feature importance, allowing us to quantify the contribution of a given feature in the predictive model. For each tree the algorithm excludes some observations, called “out-of-bag” (OOB) observations, from the original dataset. After fitting, the algorithm ranks a feature by comparing accuracy in the OOB data before and after randomizing the values of the feature in it. A larger drop in accuracy results in a higher importance score. This measure allows us to rank the features by how helpful they are in predicting a stress event for a specific individual.

We also used radial support vector machines (SVM) to classify the features from physiological signals [4, 8]. A support vector machine looks for the optimal separating hyperplane between the two classes by maximizing the margin between the classes’ closest points. The points lying on the boundaries are called support vectors with the hyperplane in the middle of the margin. When a linear separator cannot be found, which is the case for most real datasets, kernel techniques are used to project the points into a higher-dimensional space where they become linearly separable. A variety of kernels can be used including polynomial, radial basis function (RBF), and sigmoid. SVM models can also have a “hard” or “soft” margin depending on if they allow for misclassified points (soft) or not (hard). With a soft margin, which is more commonly used for real datasets since they have noise, data points on the “wrong” side of the discriminant margin are weighted down to reduce their influence. We used a soft margin due to the noisy nature of our data.

We used a radial SVM because of its good general performance in classification tasks and few number of parameters (two). The two parameters in radial SVM are cost and sigma (sometimes called gamma). Cost or cost of misclassification trades off correct classification of training examples against maximization of the decision function’s margin. The gamma or sigma parameter defines how far the influence of a single training example reaches. Because SVMs can be quite sensitive to the proper choice of parameters, the best results are obtained by checking every combination of parameters through a grid search [6]. While radial SVM does not have built-in feature importance scores, a quantitative comparison of feature importance can still be obtained using the varImp() function in *caret* [11], which computes and compares the AUC for each predictor.

### 3.2 Models Developed

For our models, we used the R *caret* package (version 6.0-85) [11], which calls the *randomForest* package (version 4.6-14) and the *kernlab* package (version 0.9-29) support vector machine. We developed a set of models for each individual. For each data set, we developed 5 random forest models based on the features of a single metric, one overall random forest model that used all 4536 features from all the metrics, 5 radial SVM models based on the features of a single metric, and one overall radial SVM model that used all 4536 features. We used the features extracted by *tsfresh* as the predictor and whether the participant was stressed or not as the binary response.

The tuning parameters were determined using cross-validated grid search, the default search process in R’s *caret* package. We set the number of trees at 500 with 10-fold cross validation repeated 3 times. For the SVM, we also used 10-fold cross validation repeated 3 times with the 90% non-validation dataset to pick the cost and sigma parameters before testing the model on the 10% validation set.

## 4. Results

When a binary classifier is applied to a new time series, it reports a probability that the time series sequence represents a high-stress state for a certain individual. A threshold can then be used to determine whether to deliver an intervention in response to this time series. At each threshold, false negatives or false positives may be generated. The trade-off between false positives and false negatives at different levels of threshold can be captured in the Receiver Operating Characteristic (ROC) plot. The area under the ROC curve (AUC) provides a measure of accuracy across all possible thresholds. The larger the AUC, the better the discrimination between the two classes. Because AUC gives a concise summary of the classifier’s performance over a range of thresholds, we used it to compare different models.

As shown in Fig. 3a, movement (ACC) is the best single-metric classifier (Avg AUC = 0.874), showing a substantially higher classification accuracy than the other single-metric classifiers (Avg AUC in the range of 0.710 − 0.796). The overall classifier (Avg AUC = 0.864, SD = 0.067) performs similarly to the ACC classifier. These two classifiers (SD of AUC: 0.070, 0.067) are also substantially more consistent across individuals than the other single-metric classifiers (SD of AUC in range 0.125-0.146). This demonstrates that these two models can predict the onset of stress 15 minutes prior accurately for various individuals, despite individual differences.

**Figure 3:**
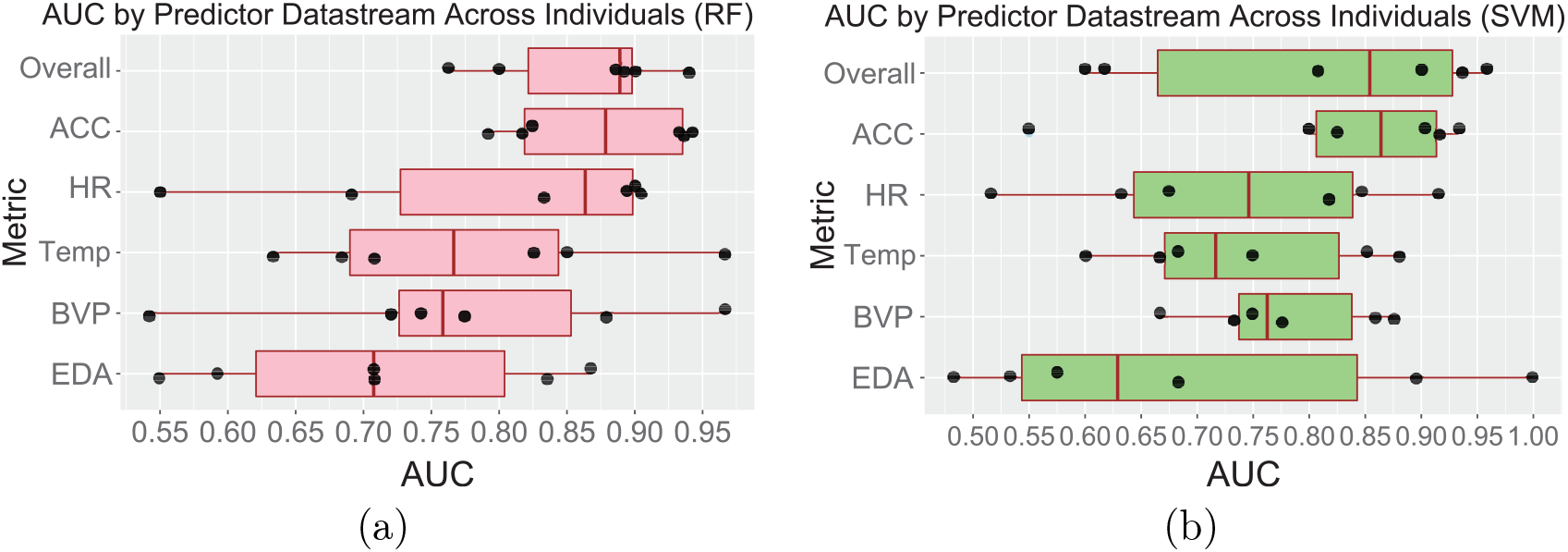
Performance comparisons (AUC) of overall and single-metric classifiers. (a) RF and (b) SVM. Each black dot is a participant.

For the SVM models, we split our data into test/train and validation sets in order to tune for the parameter C. We report AUCs from the cross validation to pick C and accuracies from the validation set. The overall SVM models had an average AUC of 0.803, which is lower than the average for the overall RF models (0.864). The SVM models also varied much more across individuals in AUC (SD of AUC = 0.160) than the RF models (SD of AUC = 0.067). As shown in Fig. 3b, the highest AUC was over 0.950 and the lowest was 0.600. We then ran the model on the validation datasets. These validation datasets had very few points (2 − 6), so we consider the accuracies we observed ranging from 0.5 to 1 to satisfactorily demonstrate that our model works on novel data. The two of the participants whose overall models had AUC ≥ 0.9 had validation accuracies of 1. The third participant’s had accuracy 0.67. The models with AUC < 0.7 had accuracies of 0.5 and the model with AUC between 0.7 and 0.9 had accuracy 0.75.

As with the RF single-metric classifiers, the ACC classifier (Avg AUC = 0.821, SD of AUC = 0.143) is significantly more accurate than the rest of the single-metric classifiers and similar to, but slightly better than, the overall model. However, as observed in the RF models, each single-metric classifier performed well on some participants and very poorly on other participants, with the rest falling somewhere in between.

In Fig. 4, we compared the two overall models for each individual using ROC curves. The overall SVM model performed well for four participants but very poorly on the other two (Individuals 1 and 6), whereas the overall RF model performed relatively well on all six participants. For Individual 2, the two models performed similarly. For Individuals 3, 4, and 5, the RF model either trailed the SVM or traced a similar curve before picking up in the right section of the curve at lower thresholds. In summary, RF has a more consistent performance than SVM across individuals and performs better than or similar to SVM in most cases.

**Figure 4:**
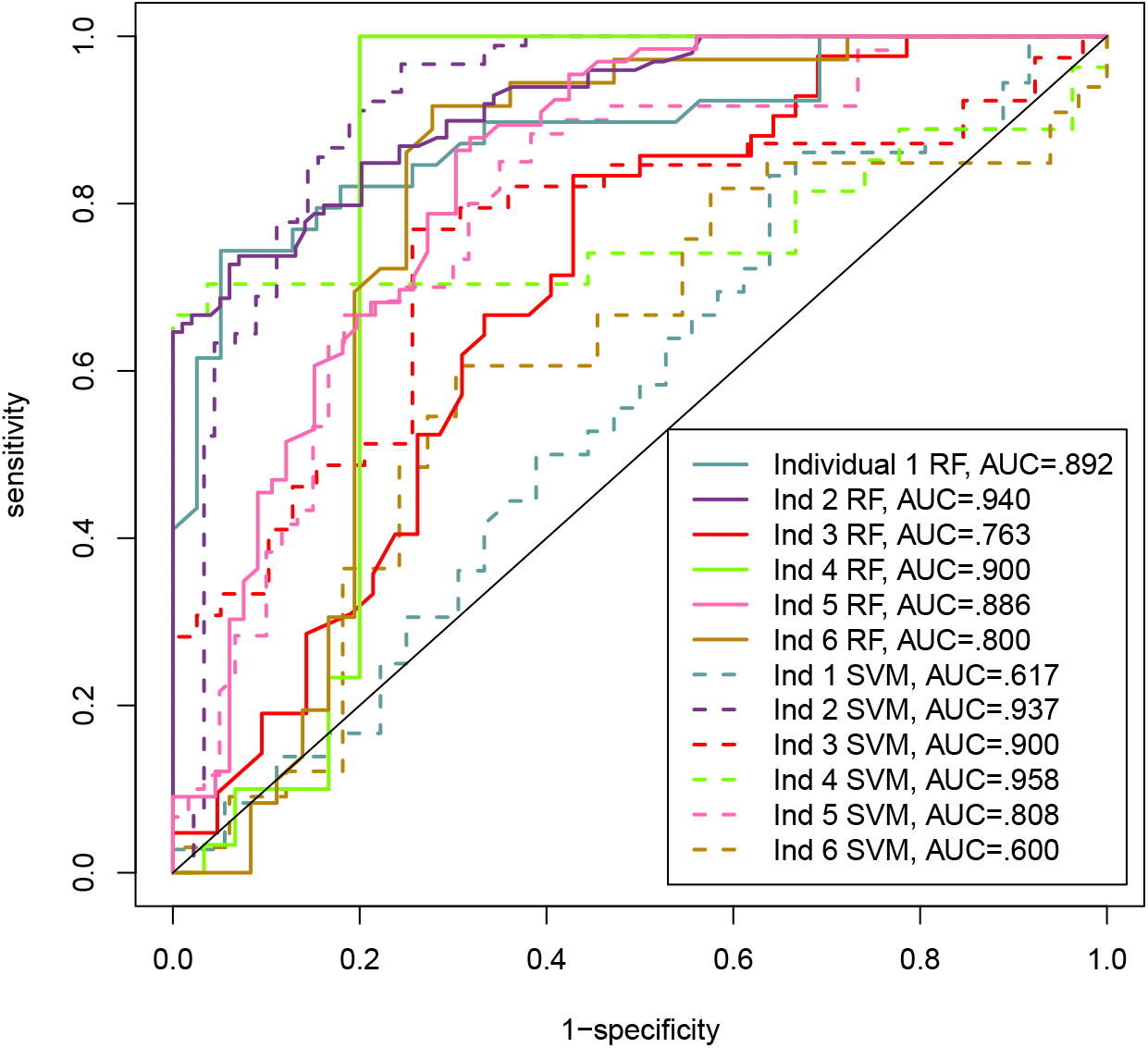
Comparison of the ROC curves of the overall RF and SVM models for each individual.

To identify predictive features, we obtained the feature importance for each feature in the overall classifier for each individual from random forest. To visualize the importance of each metric, we extracted the feature with the highest feature importance in each metric, and used it to represent the importance of the corresponding metric. Fig. 5a plots the feature importance of these representing features across individuals. We find that the representing features for HR and ACC have a much higher median feature importance than those for the other metrics. This is consistent with the observation in Fig. 3a, i.e. the ACC and HR classifiers having the highest AUC medians among all single-metric classifiers. Interestingly, we also observe a huge variation in feature importance across individuals for the same metric. A metric that is highly predictive for stress in one individual may be unimportant for the prediction in another individual.

**Figure 5:**
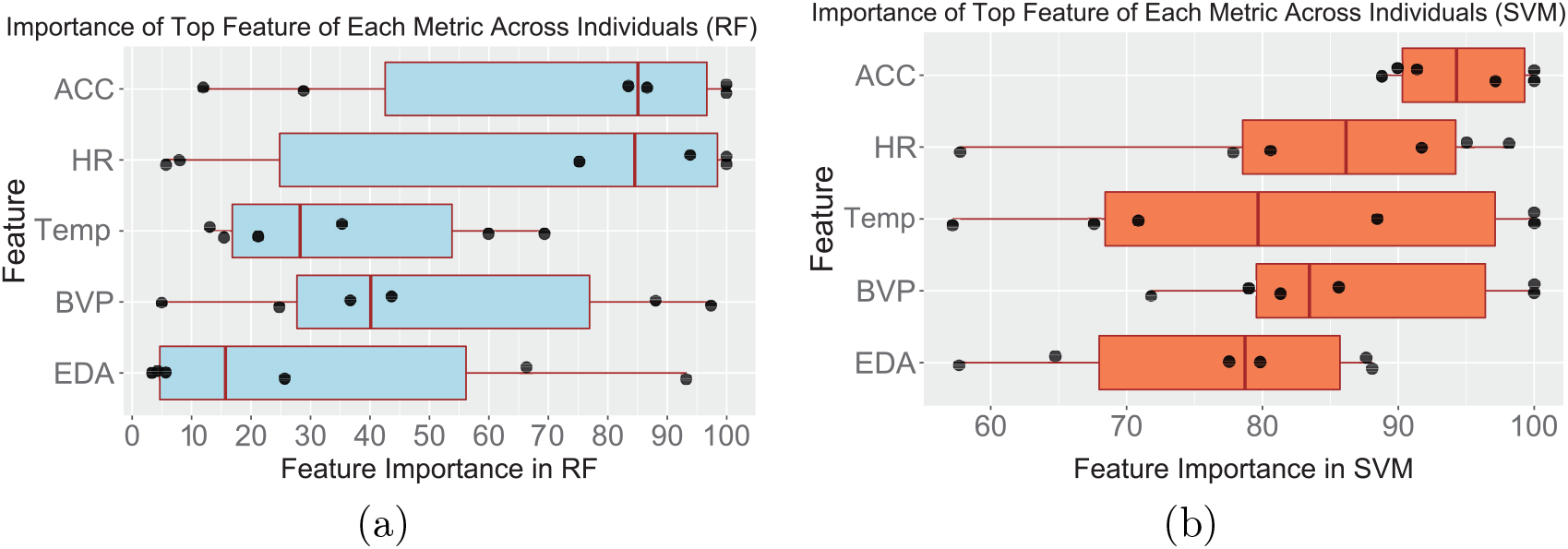
Feature importance for the most important feature of each metric type in the overall classifier. (a) RF and (b) SVM. Each black dot represents an individual.

Fig. 5b shows the feature importance of the most important feature of each metric in each overall SVM model. Similar to RF, the importance scores of the representative features for HR, Temp, BVP, and EDA in both models vary greatly across participants. However, for the SVM, ACC has a much smaller range than the other metrics, while in the RF models, ACC has a similar range to the other metrics. Based on this result, SVM appears to be more sensitive to movement-related signals of stress than RF.

We further explored the individual difference for the metrics. Fig. 6a plots the metrics that are represented in the top 10 most important features for each individual’s overall RF model. Some individuals show very strong signs in a specific metric but little in other metrics, such as heart rate (Individual 5, Fig. 6a) or ACC (Individual 1 and 5, Fig. 6a). For Individual 5, whose top 10 features consisted of nine HR features and one ACC feature, we observed that features of other metrics had either 45% importance or lower. This is consistent with the participant reports from this individual that she generally feels her heart pound rapidly when stressed and has a hard time sitting still when extremely stressed. Individual 1’s top 10 features consisted of only ACC features, indicating that she may move around a lot or fidget when nervous. ACC appears in the top 10 most important features for all the participants, further indicating that it is generally important for stress prediction. However, each individual has very different physiological signals for the onset of stress. Thus, individualized models are required for accurate prediction of stress; there is no one-size-fits-all model.

**Figure 6:**
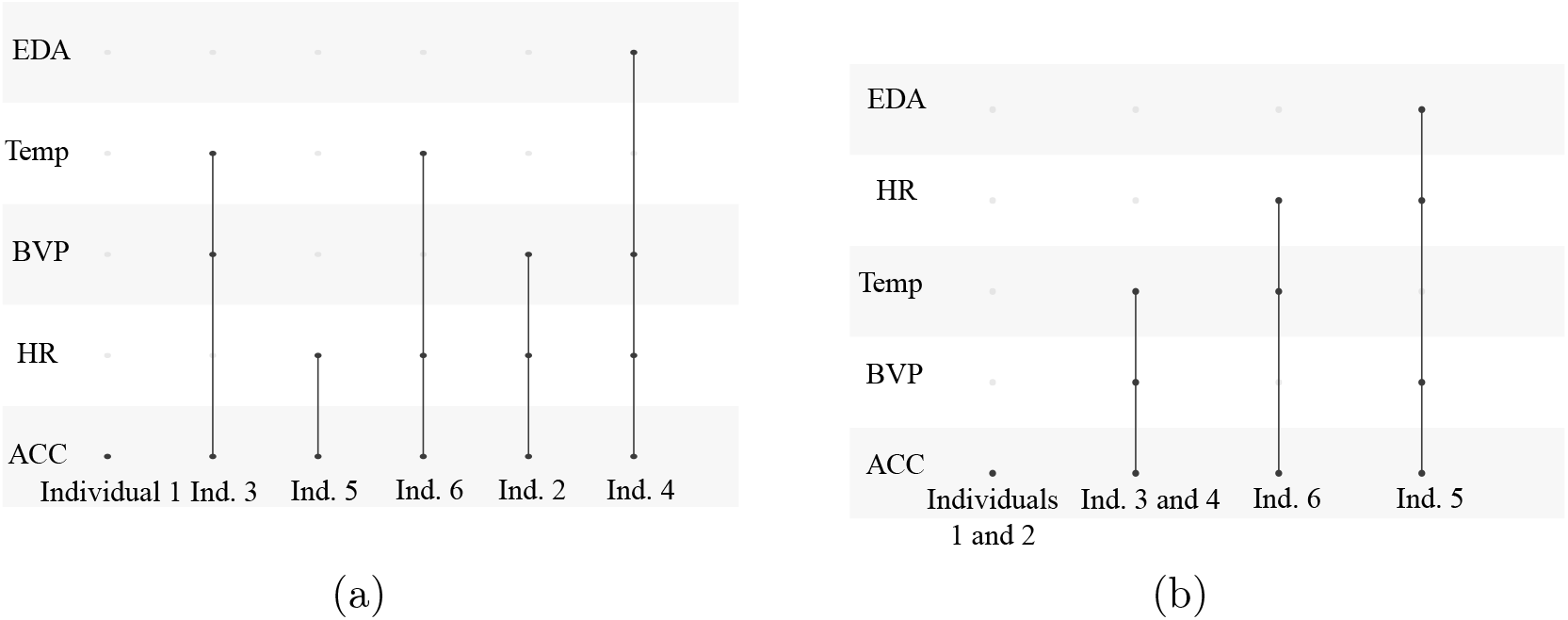
Metrics that are in the top 10 features for each individual’s overall model. (a) RF and (b) SVM. Dark points indicate a feature associated with the metric can be found in the individual’s top 10 most important features.

Next, we compared the metrics that appeared in the top 10 most important features in the two overall models. For three of the participants (Individuals 1, 3, and 6), the SVM and RF models agree with each other. Additionally, ACC appeared in the top 10 for all the participants in both models. Some changes are observed in the top 10 for Individuals 2, 4, and 5. Individual 2 went from having three metrics (ACC, HR, BVP) in the overall RF model to just one (ACC) in the overall SVM model. Individual 4 went from four (ACC, HR, BVP, EDA) to three (ACC, BVP, Temp). Individual 5 went from two (ACC, HR) to four (ACC, HR, BVP, EDA). However, changes in the metrics in the top 10 do not appear to be associated with model performance. These results indicate that the SVM and RF models pick up better on different features and that ACC is a consistently good stress predictor.

To assess the sensitivity of our RF model to the choice of non-events, we chose four different sets of non-events for Individual 5, who collected data from October through January, creating 4 different datasets. For each dataset, we trained an overall RF and four single-metric (EDA, BVP, Temp, HR) RF models. As shown in Table 1, the overall RF model performs consistently across different datasets, maintaining a high AUC in all datasets (Range: 0.827 − 0.894). In contrast, the performance of the single-metric classifiers are much more variable across datasets, depending on the choice of non-events This result demonstrates the robustness of the overall RF model, suggesting that it could be used in an application that periodically retrains with a new dataset from the user.

## 5. Conclusion and Future Considerations

We developed a predictive model for stress based on blood volume pulse, electrodermal activity, heart rate, movement, and peripheral skin temperature collected from wearable wristbands using the random forest classifier and a radial support vector machine. The random forest model has a high predictive power across individuals for predicting stress 15 minutes prior to the onset of noticeable physical symptoms. The radial support vector machine model performed better than the random forest model for two participants and had an AUC > 0.9 for three participants, but was outperformed by the random forest model in both average AUC and standard deviation. We found that movement (ACC) was the individual metric with the highest predictive power (Avg AUC = 0.874) but each person’s specific best predictors are very individualized. Despite the small sample size, the individual variability shown in our study can be extended to the general population. Therefore, every person needs a model trained specifically to their data for accurate stress prediction. Additionally, knowledge of the most important features for stress prediction in an individual could also improve their conscious understanding of their body’s signals. Because the features describe properties of the data, behaviors even more specific than simply movement or heart rate, such as finger tapping versus folding and unfolding the arms, can be pinpointed as stress indicators. Not only can this knowledge help them better notice stress on their own, but also help them develop more personalized and effective stress management practices. Additionally, paying more attention to their body’s stress signals could make teens have better mental health awareness in general. Thus, our findings could have a positive impact on teen mental health protection.

Due to the small sample size, we cannot generalize the finding that movement is particularly predictive of stress. We would also like to note that though the number of data points per person varied (20 to 66) depending on how many stress events they had (10 to 33), there was no obvious correlation between the number of data points and the model performance. It is possible that with a longer study the machine learning model performance would improve for each person, as each person has more of their own stress events for the model to learn from, resulting in less variation in the random forest and overall SVM models’ performances. However, ecological momentary assessment studies with self-reports each day generally do not run longer than 4 weeks because the participant compliance rate drops after that point. Thus, some significant changes to the data collection process will be needed in order to collect more classified target states per individual. Nevertheless, these data provide evidence that the random forests algorithm is particularly useful for training individual-specific classifiers on physiology data collected in real-life studies.

Possible future directions include a study on adolescent males or a study with a larger sample size so we can better understand general trends in teen stress indicators. We plan to improve our random forest model by developing a systematic or automated way to pick non-events on different days from the events. We also plan to try our model on time intervals farther prior to the onset of physical stress symptoms, such as an hour or half an hour. This will allow our model to pick up early indicators of stress, notify the wearer, and provide timely intervention through an app.

## Data Availability

The data is hosted in the authors' in-house server. It will be available upon the publication of the manuscript.

## Acknowledgements

CWJ is supported by a grant from the Ai4All organization.

